# Locus coeruleus intensity as a marker of early-stage Alzheimer’s disease: associations with plasma pTau_217_ and memory

**DOI:** 10.64898/2026.10.07.26364896

**Authors:** Melanie Quiring, Yuhan Shuai, Wenhui Zhao, John Paul M. Alitin, Arunima Kapoor, Allison C. Engstrom, Jillian L. Joyce, Preeti Iyengar, Trevor Lohman, Basant Lashin, Aimée Gaubert, Fatemah Shenasa, Lorena Contreras, Kevin W. Law, Farrah Ferrer, Danny J.J. Wang, Elizabeth Head, Rond Malhas, Kathleen Rodgers, S. Duke Han, Julian F. Thayer, Mark W. Bondi, Mara Mather, Daniel A. Nation

**Affiliations:** Leonard Davis School of Gerontology, University of Southern California, 3715 McClintock Ave, Los Angeles, CA 90089, USA; Memory and Aging Center, Department of Neurology, University of California, San Francisco, 675 Nelson Rising Lane, San Francisco, CA 94158, USA; Department of Psychological Science, University of California, Irvine, 4201 Social and Behavioral Sciences Gateway, Irvine, CA 92697, USA; Keck School of Medicine, University of Southern California, 1975 Zonal Ave., Los Angeles, CA 90033, USA; Stevens Neuroimaging and Informatics Institute, University of Southern California, 2025 Zonal Ave, Los Angeles, CA 90033, USA; Department of Pathology and Laboratory Medicine, University of California, Irvine, Medical Sciences D, Irvine, CA 92617, USA; Department of Neurology, University of California, Irvine, 1001 Health Sciences Road, Irvine, CA 92697, USA; Center for Innovations in Brain Science, Department of Pharmacology, University of Arizona, 1501 N. Campbell Ave., Tucson, AZ 85724-5050, USA; Department of Psychology, University of Southern California, Dornsife College of Arts and Letters, G. Mudd Building, 3620 McClintock Ave, Los Angeles, CA 90089, USA; Department of Psychiatry, University of California, San Diego, 9500 Gilman Dr, La Jolla, CA 92093, USA; Center for Affective and Autonomic Neuroscience (CAAN), Max Planck Institute for Human Development, Berlin, Germany

**Author notes:** These authors contributed equally to this work (Melanie Quiring, Yuhan Shuai). **Corresponding Author:** Daniel A. Nation, Ph.D., Leonard Davis School of Gerontology, University of Southern California, 3715 McClintock Ave, Los Angeles, CA 90089, USA.

## Abstract

**INTRODUCTION:** The locus coeruleus (LC) is one of the earliest sites of Alzheimer’s disease (AD) pathophysiological changes. LC magnetic resonance imaging (MRI) intensity may reflect effects of AD pathophysiology on the LC, providing a biomarker of early-stage AD. This study examined relationships among MRI-derived LC intensity, plasma phosphorylated tau 217 (pTau_217_), and memory in older adults without dementia.

**METHODS:** Independently living adults aged 59–89 years (*N* = 163) underwent neuropsychological assessment and T1-weighted fast spin-echo MRI. We quantified a bilateral LC contrast ratio relative to a pontine reference region, averaged across axial slices. Plasma pTau_217_ was measured using the Quanterix ALZpath p-Tau217 Advantage PLUS assay. For a subset (*n* = 127), a memory composite score was calculated.

**RESULTS:** After adjustment for age, sex, and *APOE* ɛ4 status, lower LC intensity was associated with higher plasma pTau_217_ concentrations (β = −0.17, *P* = .02). Each 1-*SD* decrease in LC intensity was associated with 53% higher odds of pTau_217_ positivity (*P* = .03). In models additionally adjusted for education, a significant LC × pTau_217_ interaction for memory (β = −0.18, *P* = .009) indicated that lower LC intensity was associated with worse memory at low (*P* = .01), but not high (*P* = .64) pTau_217_ levels.

**DISCUSSION:** These findings support MRI-derived LC intensity as a marker of early AD-related processes and suggest that LC intensity may provide complementary information not captured by plasma pTau_217_ alone. Longitudinal studies are warranted to determine how these relationships change across the early stages of AD.

## 1 | BACKGROUND

Extracellular amyloid beta (Aβ) plaque deposition and intracellular tau neurofibrillary tangle formation are hallmark features of Alzheimer’s disease (AD) that begin years or decades before clinical symptoms emerge.^1,2^ The locus coeruleus (LC) is one of the earliest sites of AD pathophysiological changes, where hyperphosphorylated tau accumulates prior to cortical tau deposition^3,4^ and the onset of cognitive symptoms.^4,5^ LC alterations are thought to contribute to disease progression in the early stages of AD^6–9^ and can be detected *in vivo* using magnetic resonance imaging (MRI), with LC intensity commonly quantified as a contrast ratio reflecting LC signal intensity relative to a nearby pontine reference region.^9,10^ Several studies examining LC intensity and tau positron emission tomography (PET) demonstrate that lower LC intensity correlates with greater AD-related tau burden,^4,11–14^ while longitudinal findings further suggest that reduced LC intensity predicts subsequent cortical tau accumulation.^14,15^ As such, MRI-derived LC intensity has been proposed as a potential biomarker of early AD-related processes and disease progression.^4,9,13–16^

Although amyloid and tau PET provide well-established *in vivo* measures of AD pathology,^17–19^ their accessibility is limited.^20,21^ Among blood-based biomarkers that offer a more scalable alternative for detecting and monitoring AD-related pathophysiological changes,^22,23^ plasma pTau_217_ is one of the most promising.^22,24^ Although pTau_217_ shows high accuracy for detecting cerebral amyloidosis during the preclinical stage of AD,^22,24,25^ it reflects early changes in soluble tau phosphorylation and release before aggregated tau is detectable with PET.^17,26,27^ Plasma pTau_217_ is increasingly incorporated into research settings,^20,24,28,29^ and several assays are now available for clinical use.^22,30,31^ Despite its increasing use as a marker of early AD pathophysiology, few studies have examined plasma pTau_217_ along with LC MRI measures in older adults, and studies to date have yielded inconsistent results. Some studies have reported no significant relationship between LC intensity and pTau_217_^32^ or a relationship limited to patients with dementia.^33^ In contrast, evidence from an adult-lifespan cohort suggests that the relationship between lower LC intensity and higher plasma pTau_217_ becomes evident around age 60 in cognitively unimpaired adults. However, the study sample included a relatively modest number of older adults.^34^ It therefore remains unclear whether MRI-derived LC intensity is associated with plasma pTau_217_ in older adults without dementia.

Understanding the relationship between LC MRI measures and plasma pTau_217_ is particularly important because LC alterations and AD pathophysiology may interact to influence memory. The LC-norepinephrine system modulates memory function and hippocampal synaptic plasticity,^8,35^ and preserved LC integrity may help maintain cognitive function in older age.^8^ Accordingly, MRI-derived measures of LC intensity are associated with better memory^36–38^ and slower rates of cognitive decline in older adults.^11,13,39^ Importantly, lower LC intensity and faster LC degeneration are associated with greater memory decline when amyloid PET burden is elevated,^4,11^ suggesting that the role of the LC in memory resilience or vulnerability may depend on underlying AD proteinopathic changes. Evidence using blood-based biomarkers is more limited. In adults aged 30–85, lower LC intensity is associated with worse global cognition at higher plasma pTau_231_ levels,^34^ but memory-specific outcomes were not examined. Subtle memory decline is observed in preclinical AD,^40,41^ and evidence suggests that memory may be affected jointly by LC degeneration and AD pathophysiology.^4,11^ Thus, investigation of the interaction between LC intensity and plasma pTau_217_ levels will help clarify whether lower LC intensity is associated with worse memory at an early stage in the AD process. The present study extends prior work connecting LC intensity with AD pathophysiology and memory by examining these relationships using plasma pTau_217_ in older adults with unimpaired or mildly impaired cognition.

## 2 | METHODS

### 2.1 | Participants

Participants were independently living adults aged 55–90 enrolled in the Vascular Senescence and Cognition (VaSC) Study at the University of Southern California (USC) and the University of California, Irvine (UCI). Participants were recruited through word-of-mouth, flyers, community events, or community participant registries. Exclusion criteria were a history of clinical stroke, dementia, moderate or severe traumatic brain injury, neurological or psychiatric disorders that could impair cognitive function, or MRI contraindication. Upon completion of MRI, each participant’s T2-weighted scan was reviewed and abnormalities were referred to a board-certified neuroradiologist for further evaluation. Participants with confirmed gross abnormalities (e.g., silent infarctions, frontal gliosis, and lobar cavernoma) were excluded. Among the remaining VaSC participants, 169 had valid LC imaging and plasma pTau_217_ data. This study was approved by the USC and UCI Institutional Review Boards. All participants provided written informed consent and were treated in accordance with the Declaration of Helsinki.

### 2.2 | MRI Acquisition and Processing

Participants underwent brain MRI on a 3T Siemens MAGNETOM Prisma scanner. High-resolution T1-weighted structural images were acquired using a magnetization-prepared rapid acquisition gradient-echo (MPRAGE) sequence (TR = 2300 ms; TE = 2.98 ms; TI = 900 ms; flip angle = 9°; field of view = 256 mm; voxel size = 1.0 × 1.0 × 1.2 mm³). LC imaging was performed using a two-dimensional T1-weighted fast spin-echo (T1-FSE) sequence, as previously described,^42^ acquired perpendicular to the brainstem and positioned to encompass the rostrocaudal extent of the pons. Acquisition parameters were TR = 750 ms, TE = 12 ms, flip angle = 120°, bandwidth = 287 Hz/pixel, voxel size = 0.43 × 0.43 × 2.5 mm, slice gap = 1 mm, and 11 axial slices.

T1-FSE images were processed using a previously developed semi-automated LC delineation and registration procedure.^38^ FSE images were coregistered to each participant’s native-space MPRAGE image using antsRegistrationSyNQuick.sh in Advanced Normalization Tools (ANTs; version 2.5.0).^43^ Separate study-specific MPRAGE and FSE templates were subsequently constructed using antsMultivariateTemplateConstruction.sh from the native-space MPRAGE images and the corresponding MPRAGE-aligned FSE images, respectively.

Registration between the study-specific MPRAGE and FSE templates, and between the study-specific MPRAGE template and the MNI–ICBM 152 linear template at 0.5-mm isotropic resolution, was performed using antsRegistration. The MNI-space high-confidence LC meta-mask and central pontine reference mask described by Dahl et al.^12^ were then transformed into the study-specific FSE-template space using inverse transformations. Participant FSE images resampled into the study-specific FSE-template space were used for LC signal-intensity extraction. For each axial slice containing matched LC and pontine reference regions, the maximum signal intensity within the LC mask and the maximum signal intensity within the pontine reference mask were extracted. A slice-specific LC contrast ratio was then calculated separately for the left and right LC as:

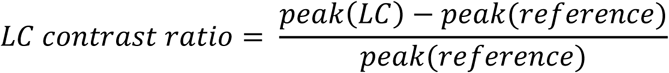

where *peak(LC)* and *peak(reference)* represent the maximum signal intensities within the LC and pontine reference regions, respectively, for that slice.^38,42,44^ This procedure was repeated across matched slices, and the resulting slice-wise contrast ratios were averaged across slices to obtain mean left and right LC contrast ratios for each participant.

### 2.3 | Neuropsychological Assessment

Participants underwent a comprehensive neuropsychological test battery within 4.6 months of MRI (mean interval = 1.1 month). Cognitive status was determined as cognitively unimpaired (CU) or probable MCI using the Jak-Bondi criteria^45^ based on ten neuropsychological test scores assessing global cognition (Mattis Dementia Rating Scale-2 or Montreal Cognitive Assessment), memory (Craft Story 21 Delayed Recall or Logical Memory II Delayed Recall, California Verbal Learning Test [CVLT] Long Delay Free Recall or Rey Auditory Verbal Learning Test [RAVLT] Delayed Recall, and CVLT Recognition Discriminability or RAVLT Recognition Percent Correct), executive function (Trail Making Test Part A, Trail Making Test Part B, and either the Stroop Color-Word Interference Test, Delis-Kaplan Executive Function System [D-KEFS] Color-Word Interference Test, or Digit Span Backward), and language (FAS Letter Fluency, Animal Semantic Fluency, and Multilingual Naming Test or Boston Naming Test). Due to differences in test batteries across participants, analyses of memory were restricted to a subset of 127 participants who completed the Logical Memory and RAVLT measures used to derive the memory composite. The memory composite was calculated using raw scores from Logical Memory II Delayed Recall, RAVLT total learning across Trials 1–5, and RAVLT Delayed Recall. Individual test scores were standardized (*z*-scored) and averaged to create the composite score, with higher values reflecting better memory performance.

### 2.4 | pTau_217_ Quantification

Blood samples were collected at either the MRI or neuropsychological assessment visit after overnight fasting. Plasma pTau_217_ concentrations were measured using the ultrasensitive Single Molecule Array (Simoa) platform on the Quanterix HD-X Analyzer. Blood samples were collected in K2EDTA tubes and centrifuged at 2000 × g for 10 minutes at 4°C. The isolated plasma was transferred and were aliquoted into cryovials. The plasma aliquots were transferred into a −80 °C freezer for storage until required for analyses. For the assay, plasma samples were thawed on ice, centrifuged to remove residual cellular debris, and processed according to the manufacturers’ protocol. Plasma pTau_217_ was quantified using the Simoa ALZpath p-Tau 217 Advantage PLUS Kit (Item No. 104570). The assay uses a bead-based two-step sandwich immunoassay in which pTau_217_ specific capture antibodies are coupled to paramagnetic beads, and detector antibodies are linked to an enzyme label. Following sequential incubation and wash steps, immunocomplexes were distributed into a microwell array for digital single-molecule detection. Chemiluminescent signal was then measured by the HD-X Analyzer to determine plasma pTau_217_ concentrations. For secondary analyses, participants were classified as pTau_217_ positive or negative using a previously established cutoff optimized for the detection of cerebral amyloidosis on Aβ PET imaging (0.42 pg/mL),^22^ with positivity indicated by values > 0.42.

### 2.5 | Covariates

Age, sex, and years of education were self-reported during the study interview. Cognitive status was classified as CU or MCI as described above. *APOE* genotyping was preformed on genomic DNA isolated from blood using one of two PCR-based methods: a TaqMan SNP genotyping assay targeting rs429358 and rs7412^46^ or PCR followed by restriction enzyme digestion and gel electrophoresis.^47^ *APOE* ε4 carrier status was defined by the presence of at least one ε4 allele.

### 2.6 | Analytic Plan

All analyses were conducted in R (version 4.6.1). Statistical significance was defined as *P* < .05 (two-tailed). Mean left and right LC contrast ratios were averaged to derive a bilateral LC intensity value for each participant. Prior to statistical analyses, bilateral LC intensity values were screened for outliers, and values exceeding ±3 standard deviations from the sample mean were excluded. A linear regression model tested the association between LC intensity and plasma pTau_217_ concentration, adjusting for age, sex, and *APOE* ε4 carrier status (*APOE* ε4 positive or negative). As a secondary analysis, logistic regression was used to examine whether LC intensity was associated with plasma pTau_217_ positivity, adjusting for the same covariates. LC intensity was standardized (z-scored) and reverse coded for this analysis so that the odds ratio represents the change in odds of pTau_217_ positivity associated with a 1-*SD* decrease in LC-CR.

In a subset of participants (*n* = 127) with complete data for the memory composite, a linear regression model tested the LC intensity × pTau_217_ interaction on memory composite scores, adjusting for age, sex, education, and *APOE* ε4 carrier status. LC intensity and pTau_217_ were mean-centered prior to constructing the interaction term to improve interpretability. Significant interactions were probed using simple slopes analyses^48^ to estimate the association between LC intensity and memory at low (–1 *SD*), mean, and high levels (+1 *SD*) of pTau_217._ Johnson–Neyman analyses were used to determine the range of pTau_217_ values at which LC intensity is significantly associated with memory, with pTau_217_ thresholds reported in raw units. As a secondary analysis, the LC intensity × pTau_217_ interaction on memory was repeated using binary pTau_217_ status (negative or positive) in place of continuous pTau_217_.

To account for potential differences in plasma pTau217 levels associated with cognitive status, significant results were followed by sensitivity analyses additionally adjusted for cognitive status (CU or MCI).

## 3 | RESULTS

Of the 169 participants with valid plasma pTau_217_ and LC imaging data, two participants with extreme negative LC intensity values were excluded upon outlier screening. Of the remaining 167 participants, four did not have valid *APOE* genotype data and were excluded from analyses, resulting in a final analytic sample of 163 participants. Participant characteristics are shown in Table 1. The majority of participants (77.9%) identified as non-Hispanic White, followed by 12.9% as Asian, 5.5% as Black or African American, and 3.7% as Hispanic. A subset of participants (*n* = 127) who completed the neuropsychological tests comprising the memory composite were included in analyses examining the interactive effects of LC intensity and plasma pTau_217_ on memory performance. Participants included in the memory subset did not differ from those excluded from the memory analyses on education, plasma pTau_217_, LC intensity, sex, *APOE* ε4 carrier status, MCI status, or pTau_217_ positivity (all *P* > .05), but were younger (69.53 ± 7.28 vs. 73.17 ± 6.92 years, *P* = .008).

**TABLE 1.**
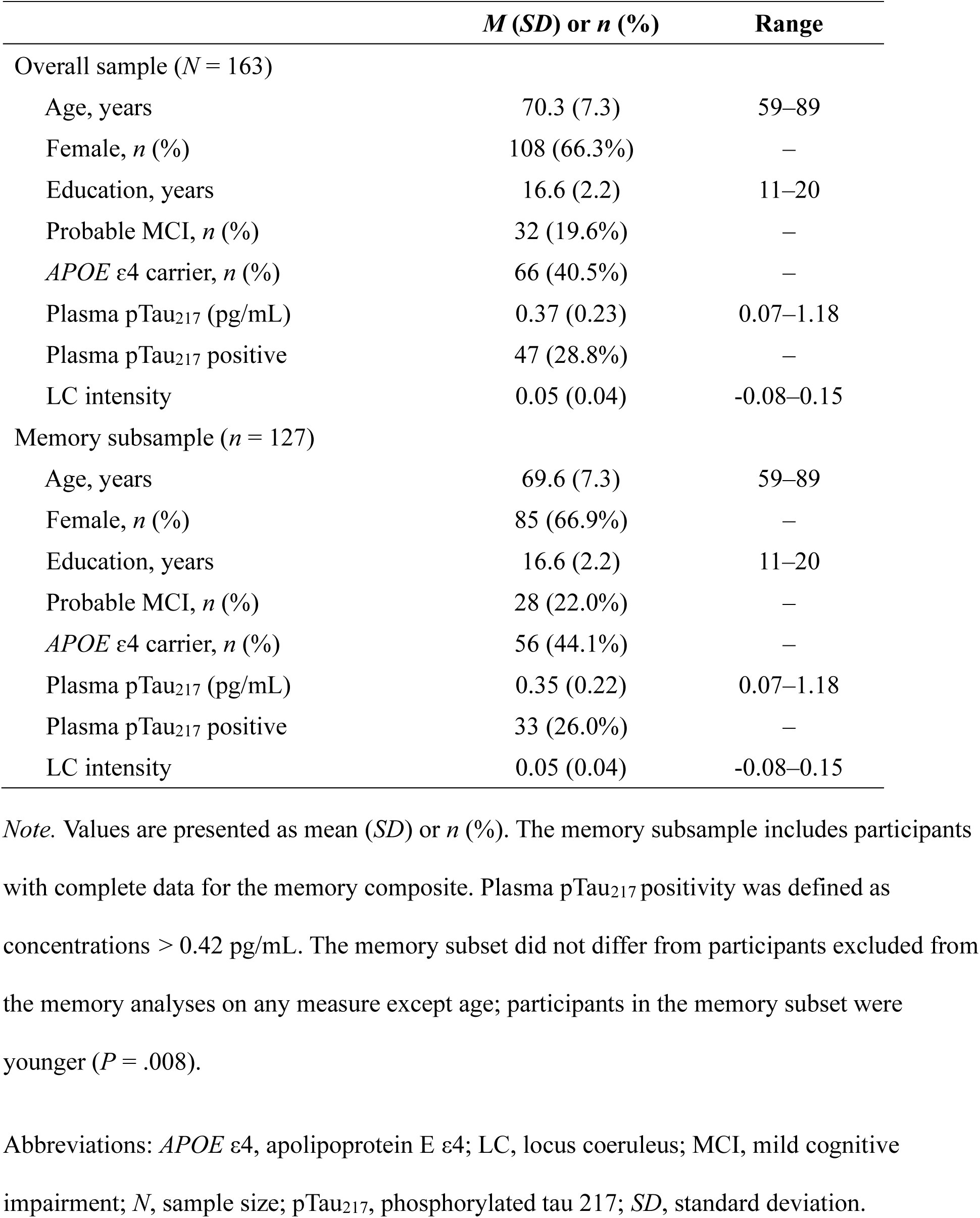
Participant characteristics.

### 3.1 | LC intensity relationship with plasma pTau_217_

Results from the multiple linear regression model examining the association between LC intensity and plasma pTau_217_ are presented in Table 2 and illustrated in Figure 1. After adjusting for age, sex, and *APOE* ɛ4 carrier status, lower LC-CR is associated with higher plasma pTau_217_ levels (standardized β = –.17, *P* = .02). Consistent with this finding, in a secondary logistic regression analysis, each 1-*SD* decrease in LC intensity was associated with 53% higher odds of plasma pTau_217_ positivity (OR = 1.53, 95% CI [1.05, 2.31], *P* = .033; Supplemental Table S1). In sensitivity analyses additionally adjusting for cognitive status, both associations remain significant (plasma pTau_217_ concentration: *b* = −1.01, *SE* = 0.46, *P* = .029; pTau_217_ positivity: OR = 1.50, 95% CI [1.02, 2.27], *P* = .044; Supplemental Tables S2 and S3).

**Figure 1.**
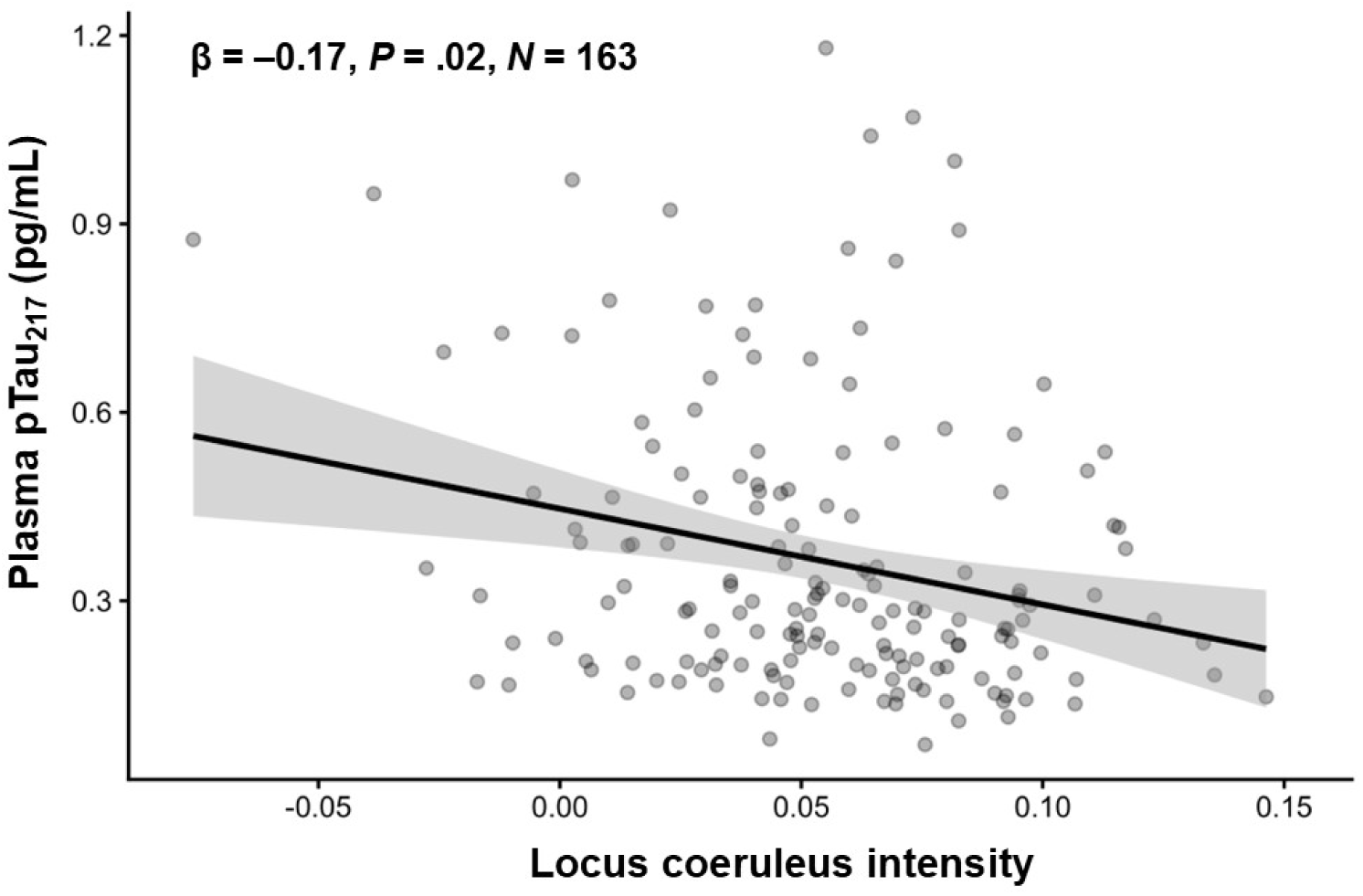
Lower LC intensity is associated with higher plasma pTau_217_ levels after adjustment for age, sex, and *APOE* ε4 carrier status. The solid line represents the fitted linear regression line, and the shaded band represents the 95% confidence interval. The standardized regression coefficient (β) is displayed in the figure. Abbreviations: *APOE* ε4, apolipoprotein E ε4; β, standardized regression coefficient; LC, locus coeruleus; MCI, mild cognitive impairment; *N*, sample size; pTau_217_, phosphorylated tau 217.

**TABLE 2.** Results from multiple linear regression model examining the association between locus coeruleus intensity and plasma pTau_217_.

|  | *b* | *SE* | 95% CI | *P* |
| --- | --- | --- | --- | --- |
| Intercept | -0.43 | 0.16 |  |  |
| LC intensity | -1.05 | 0.45 | -1.94, -0.16 | **.022** |
| Age | 0.01 | 0.002 | 0.01, 0.02 | < .001 |
| Sex | 0.01 | 0.03 | -0.06, 0.07 | .872 |
| *APOE* ε4 carrier status | 0.14 | 0.03 | 0.07, 0.20 | < .001 |

*Note.* Unstandardized estimates, standard errors, and 95% CIs are presented. *APOE* ε4 carrier status was dummy-coded such that 0 = noncarrier and 1 = carrier and sex was dummy-coded such that 0 = male and 1 = female.
Abbreviations: *APOE* ε4, apolipoprotein E ε4; *b*, unstandardized regression coefficient; CI, confidence interval; LC, locus coeruleus; pTau<sub>217</sub>, phosphorylated tau 217; *SE*, standard error.

### 3.2 | LC intensity and plasma pTau_217_ interaction on memory

Results from the multiple linear regression model examining whether plasma pTau_217_ modifies the association between LC intensity and memory are presented in Table 3 and illustrated in Figure 2. A significant LC intensity × plasma pTau_217_ interaction (standardized β = –.18, *P* = .009) indicated that the relationship between LC intensity and memory varies according to plasma pTau_217_ level. Simple slopes analyses show that at low plasma pTau_217_ levels (−1 *SD*), lower LC intensity is associated with worse memory (*b* = 7.43, *SE* = 2.81, 95% CI [1.86, 13.00], *P* = .009). The association between LC intensity and memory is not significant at mean (*b* = 3.17, *SE* = 2.02, 95% CI [–0.83, 7.18], *P* = .120) or high plasma pTau_217_ levels (+1 *SD*; *b* = −1.09, *SE* = 2.34, 95% CI [−5.72, 3.54], *P* = .636). Johnson–Neyman analyses indicate that lower LC intensity is associated with worse memory at plasma pTau_217_ concentrations below 0.31 pg/mL, encompassing 74 participants (58.3% of the analytic sample). An additional region of significance above 1.03 pg/mL included only two participants and was therefore not interpreted. Results were similar when pTau_217_ was dichotomized using the established 0.42 pg/mL cutoff, with a significant LC intensity × pTau_217_ status interaction (*b* = −10.21, *SE* = 4.11, *P* = .014; Supplemental Table S4). Lower LC intensity is associated with worse memory among pTau_217_-negative participants (*b* = 5.75, *SE* = 2.41, *P* = .019), but not among those who are pTau_217_-positive (*b* = −4.46, *SE* = 3.38, *P* = .19). These findings indicate that the relationship between lower LC intensity and worse memory is most evident at lower levels of plasma pTau_217_ and is attenuated as pTau_217_ levels increase. In sensitivity analyses additionally adjusting for cognitive status, both interactions remain significant (continuous pTau_217_: *b* = −18.73, *SE* = 6.00, 95% CI [−30.63, −6.84], *P* = .002; binary pTau_217_ status: *b* = −8.81, *SE* = 3.41, 95% CI [−15.56, −2.06], *P* = .011; Supplemental Tables S5 and S6).

**Figure 2.**
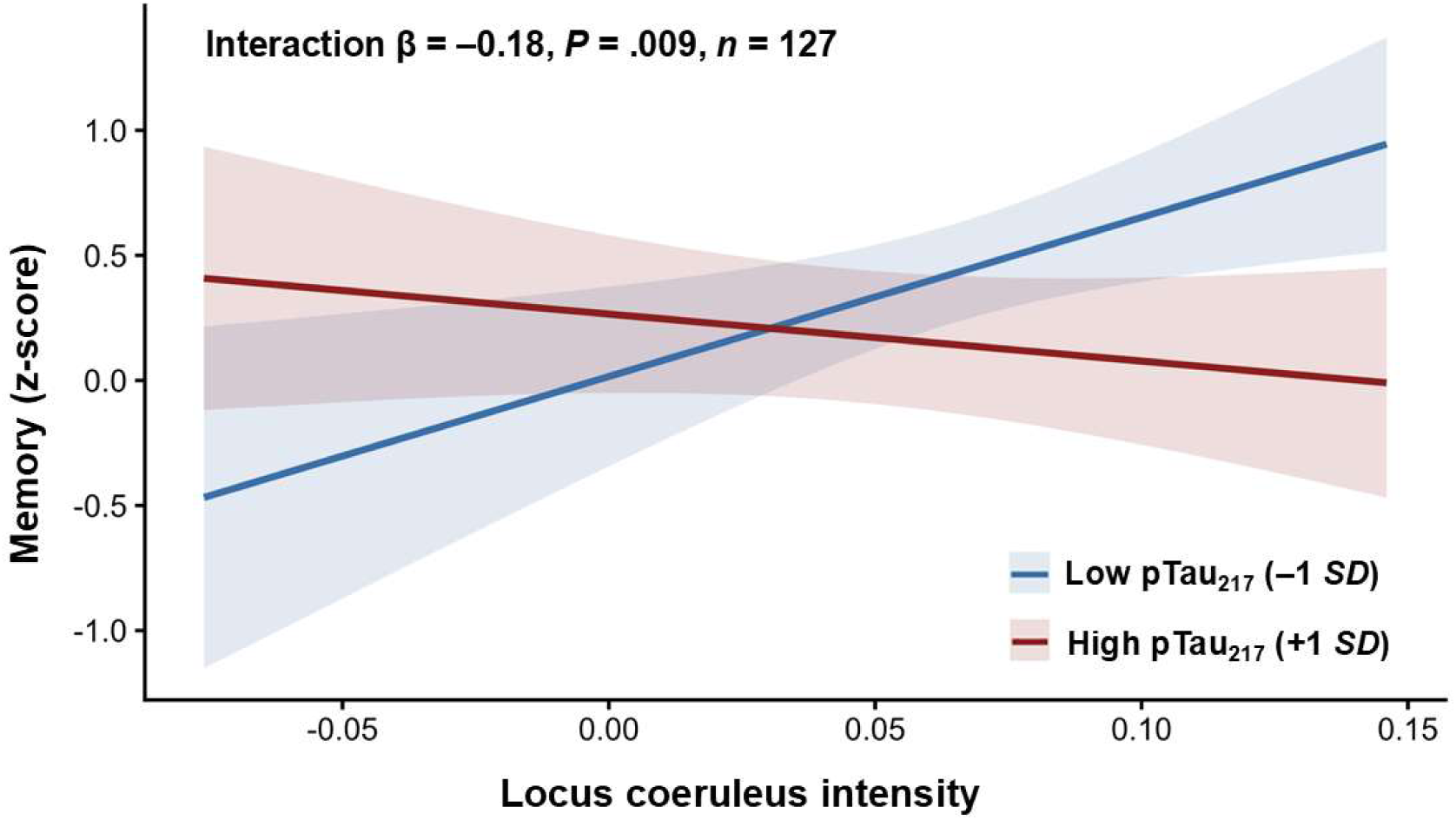
Plasma pTau_217_ modifies the association between LC intensity and memory, such that lower LC intensity is associated with worse memory at low, but not high, pTau_217_ levels. Lines represent model-estimated associations at low (−1 *SD*) and high (+1 *SD*) plasma pTau_217_ levels, and shaded bands represent 95% confidence intervals. The standardized interaction coefficient (β) is displayed in the figure. Abbreviations: *APOE* ε4, apolipoprotein E ε4; β, standardized regression coefficient; LC, locus coeruleus; MCI, mild cognitive impairment; *n*, sample size; pTau_217_, phosphorylated tau 217; *SD*, standard deviation.

**TABLE 3.**
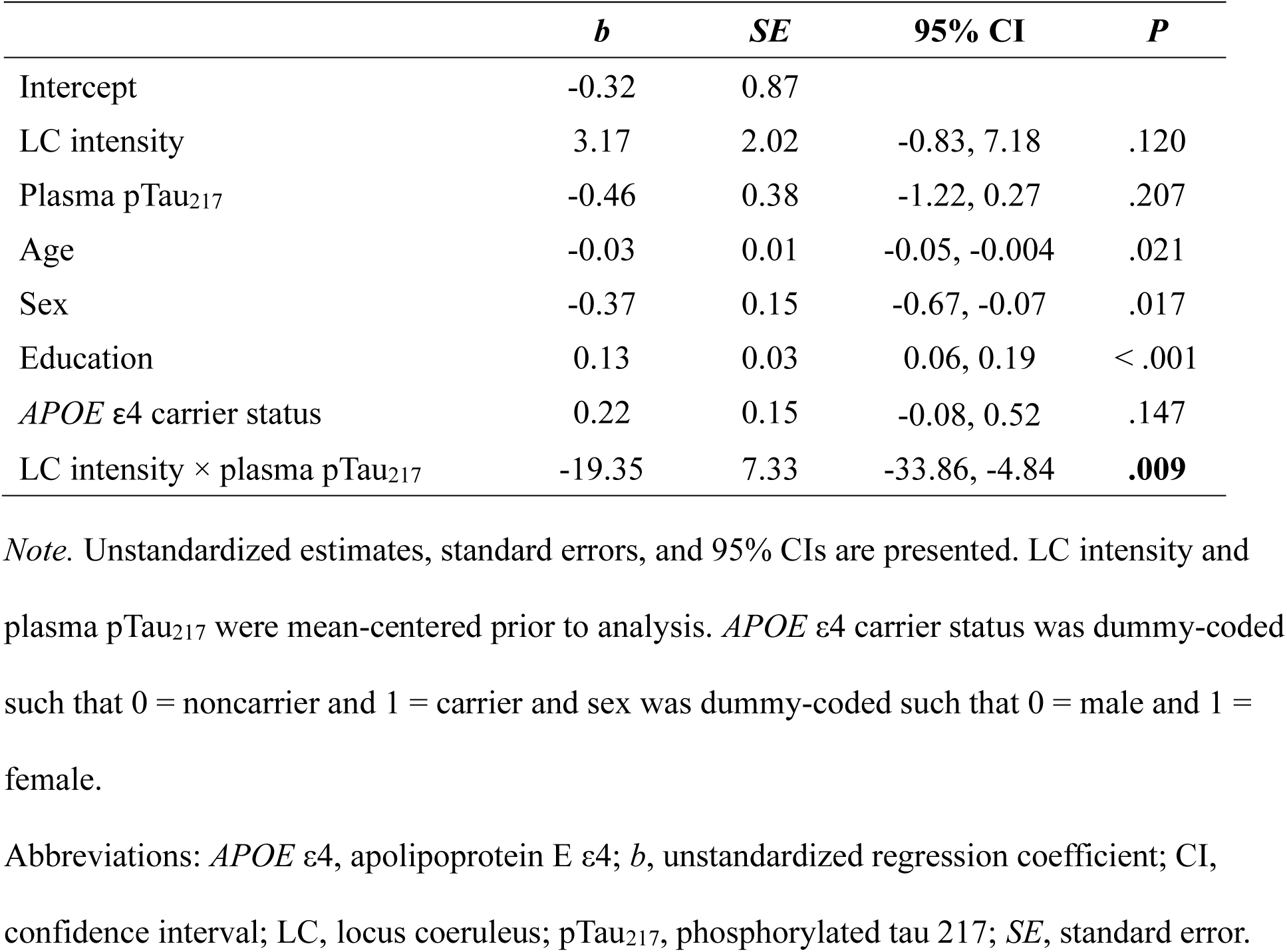
Results from multiple linear regression model examining the interaction between locus coeruleus intensity and plasma pTau_217_ in relation to memory scores.

|  | *b* | *SE* | 95% CI | *P* |
| --- | --- | --- | --- | --- |
| Intercept | -0.32 | 0.87 |  |  |
| LC intensity | 3.17 | 2.02 | -0.83, 7.18 | .120 |
| Plasma pTau<sub>217</sub> | -0.46 | 0.38 | -1.22, 0.27 | .207 |
| Age | -0.03 | 0.01 | -0.05, -0.004 | .021 |
| Sex | -0.37 | 0.15 | -0.67, -0.07 | .017 |
| Education | 0.13 | 0.03 | 0.06, 0.19 | < .001 |
| *APOE* ε4 carrier status | 0.22 | 0.15 | -0.08, 0.52 | .147 |
| LC intensity × plasma pTau<sub>217</sub> | -19.35 | 7.33 | -33.86, -4.84 | **.009** |

*Note.* Unstandardized estimates, standard errors, and 95% CIs are presented. LC intensity and plasma pTau<sub>217</sub> were mean-centered prior to analysis. *APOE* ε4 carrier status was dummy-coded such that 0 = noncarrier and 1 = carrier and sex was dummy-coded such that 0 = male and 1 = female.
Abbreviations: *APOE* ε4, apolipoprotein E ε4; *b*, unstandardized regression coefficient; CI, confidence interval; LC, locus coeruleus; pTau<sub>217</sub>, phosphorylated tau 217; *SE*, standard error.

## 4 | DISCUSSION

The findings of the present study contribute to our understanding of the relationship between LC intensity, early AD pathophysiological changes reflected by plasma pTau_217_, and memory in older adults. We first examined the association between MRI-derived LC intensity and plasma pTau_217_, hypothesizing that lower LC intensity would be associated with higher plasma pTau_217_ levels. Consistent with neuropathological evidence that the LC is an early site of AD-related tau abnormalities,^3–5^ our results indicate that nondemented older adults with less LC intensity show higher plasma pTau_217_ concentrations. This supports the potential utility of LC intensity as a marker of early AD processes and raises the possibility that LC intensity may provide information about memory-related vulnerability even before plasma pTau_217_ concentrations become abnormal. Therefore, we next examined whether the relationship between LC intensity and memory is more evident at lower or higher pTau_217_ levels, as prior work suggests that LC-related memory effects may differ according to the stage and burden of AD-related pathology.^4,11,49^ Our findings demonstrate that among older adults with unimpaired or mildly impaired cognition, less LC intensity is associated with worse memory at relatively low or subthreshold pTau_217_ levels, whereas LC intensity is not related to memory performance at higher pTau_217_ levels. Together, these results suggest that lower LC intensity corresponds with early AD-related changes in soluble tau phosphorylation, and that LC intensity may also add clinically relevant information about early memory-related processes not captured by plasma pTau_217_ alone.

Prior studies examining LC intensity in relation to AD biomarkers have primarily relied on tau PET, with several studies demonstrating that lower LC intensity tracks with cortical tau deposition in older adults without dementia^4,11,13,15^ and in autosomal dominant AD cohorts.^12,14^ However, cortical tau deposition visible on tau PET imaging is a relatively late, “Core 2” biomarker of AD that becomes abnormal later in the evolution of AD and is more closely linked with the onset of symptoms, rather than prior to symptom onset (which defines a Core 1 biomarker).^17^ Less research has focused on the relationship between LC intensity and earlier, Core 1 T_1_ blood-based AD biomarkers, such as plasma pTau_217_, that are rapidly being incorporated into clinical practice.^22,30,31^ Findings to date have also been mixed, leaving uncertainty regarding the relationship between LC intensity and plasma pTau_217_ levels in older adults without dementia.^32–34^ Importantly, previous studies examined this relationship in adult-lifespan samples^34^ or clinically mixed samples,^32,33^ with relatively modest numbers of nondemented older adults, the primary population being targeted for primary and secondary AD prevention. By identifying an association between LC intensity and plasma pTau_217_ concentrations in a larger sample of older adults without dementia, the present findings help resolve this uncertainty within the population most relevant to early AD risk detection.

Neuropathological studies identify the LC as one of the first brain regions to show abnormal tau in preclinical AD,^3–5^ with tau misfolding and seeding hypothesized to occur in the LC before propagating along its axons to medial temporal and cortical regions.^6,7,50^ The relationship between LC intensity and plasma pTau_217_ may be particularly salient during the early stages of AD since both measures capture biological changes that precede fibrillar tau accumulation and neurodegeneration. The revised Alzheimer’s Association criteria classify plasma pTau_217_ as a Core 1 T_1_ biomarker of phosphorylated and secreted AD tau that becomes abnormal around the same time as amyloid PET.^17^ Plasma pTau_217_ is thought to reflect early Aβ-related changes in soluble tau^17,20,26,51^ and is one of the earliest tau species to become abnormal across the AD continuum.^20^ Similarly, tau accumulation within the LC occurs well before appreciable LC degeneration^6,52^ and is accompanied by alterations in neuronal morphology and function.^52^ LC MRI contrast is thought to reflect the structural integrity of neuromelanin-containing LC neurons and may be sensitive to early cellular changes preceding neuronal loss,^10,11^ although the precise biological basis of LC MRI intensity remains debated.^9,53^ The relationship between lower LC intensity and higher plasma pTau_217_ observed in the present study may represent early interrelated changes within the LC-noradrenergic system and soluble tau pre-tangle pathophysiology.

In the present study, lower LC intensity relates to worse memory when pTau_217_ concentrations are relatively low or below the established cutoff associated with Aβ PET positivity, suggesting that LC-related changes may be most relevant to memory very early in the AD process. These findings are conceptually consistent with evidence from a cerebrospinal fluid biomarker study showing that lower LC metabolism is associated with faster memory decline during the early (Aβ-positive/tau-negative), but not later (Aβ-positive/tau-positive) stages of preclinical AD.^49^ The present findings demonstrate a similar pattern using MRI-derived LC intensity and suggest that LC structural intensity may also be relevant to memory function when plasma pTau_217_ concentrations remain relatively low. The pattern observed here contrasts with longitudinal PET studies showing that lower LC intensity or greater LC degeneration are associated with faster memory decline as Aβ burden increases.^4,11^ The discrepancy between the present cross-sectional findings with plasma pTau217 and these prior longitudinal Aβ PET findings may reflect differences in the biomarkers examined or stage-dependent relationships between the LC, AD proteinopathy, and memory. There is also some evidence suggesting nonlinear trajectories in LC alterations across the course of disease,^49,52^ which could account for some discrepancies in the literature. Nevertheless, the present study adds to evidence that the relationship between the LC and memory varies across early stages of AD-related pathophysiological change and suggests that MRI-derived LC intensity may be useful for characterizing early AD-related vulnerability, even before plasma pTau_217_ becomes substantially elevated.

The limitations of the present study should be considered in light of several strengths. Plasma pTau_217_ is an accessible and increasingly used biomarker that is sensitive to the earliest AD-related tau changes.^17,20^ The relatively large sample of older adults without dementia allowed us to examine relationships among LC intensity, plasma pTau_217_, and memory during early stages of AD-related biological change. Findings were robust across analyses of continuous pTau_217_ concentrations and when using an established cutpoint for pTau_217_ positivity,^22^ as well as after adjustment for cognitive status. However, the relatively small number of participants with probable MCI prevented adequately powered comparisons between unimpaired and mildly impaired participants. Although relationships between LC intensity, AD biomarkers, and cognition have been examined in patients with AD dementia,^32,33^ stage-specific relationships during MCI remain less well characterized. The spatial resolution of the LC acquisition also limited examination of anatomical specificity along the rostrocaudal extent of the nucleus, which may be important given evidence that rostral LC intensity is particularly relevant to memory and early AD-related tau changes.^5,34,38^ Finally, the cross-sectional design precludes conclusions regarding temporal ordering or directionality among LC intensity, plasma pTau_217_, and memory. Longitudinal studies incorporating repeated measures of LC intensity, plasma biomarkers, and cognition are warranted to determine how these relationships interact across the early stages of AD.

## Data Availability

The data that support the findings of this study are available upon reasonable request from the corresponding author, D.A.N., subject to institutional policies.

## Acknowledgements

This research was supported by the National Institutes of Health under grants R01AG064228, R01AG082073, P01AG052350, P30AG066530, and P30AG094848. Melanie Quiring was additionally supported by T32AG000037, S. Duke Han was additionally supported by K24AG081325, Arunima Kapoor was additionally supported by American Heart Association 23PRE1014192, and Jillian L. Joyce was additionally supported by American Heart Association 26PRE1561693.

## Non-standard abbreviations

ANTs: Advanced Normalization Tools;
CVLT: California Verbal Learning Test;
LC: locus coeruleus;
LC-CR: locus coeruleus contrast ratio;
MCI: mild cognitive impairment;
RAVLT: Rey Auditory Verbal Learning Test;
T1-FSE: T1-weighted fast spin echo.

## Notes

### Competing Interest Statement

Kathleen Rodgers is a consultant for NeuTherapeutics; this relationship is unrelated to the present work. All other authors declare no conflicts of interest.

### Author Declarations

Institutional Review Board of University of Southern California approved this work (HS-14-00784) Institutional Review Board of University of California, Irvine approved this work(HS-2019-5324)

## REFERENCES

1. Jack CR, Knopman DS, Jagust WJ, et al. Hypothetical model of dynamic biomarkers of the Alzheimer’s pathological cascade. Lancet Neurol. 2010;9(1):119–128. doi:10.1016/S1474-4422(09)70299-6

2. Villemagne VL, Burnham S, Bourgeat P, et al. Amyloid β deposition, neurodegeneration, and cognitive decline in sporadic Alzheimer’s disease: a prospective cohort study. Lancet Neurol. 2013;12(4):357–367. doi:10.1016/S1474-4422(13)70044-9

3. Braak H, Thal DR, Ghebremedhin E, Del Tredici K. Stages of the pathologic process in Alzheimer disease: age categories from 1 to 100 years. J Neuropathol Exp Neurol. 2011;70(11):960–969. doi:10.1097/NEN.0b013e318232a379

4. Jacobs HIL, Becker JA, Kwong K, et al. In vivo and neuropathology data support locus coeruleus integrity as indicator of Alzheimer’s disease pathology and cognitive decline. Sci Transl Med. 2021;13(612):eabj2511. doi:10.1126/scitranslmed.abj2511

5. Hary AT, Chadha S, Mercaldo N, et al. Locus coeruleus tau validates and informs high-resolution MRI in aging and at earliest Alzheimer’s pathology stages. Acta Neuropathol Commun. 2025;13(1):44. doi:10.1186/s40478-025-01957-6

6. Satoh A, Iijima KM. Roles of tau pathology in the locus coeruleus (LC) in age-associated pathophysiology and Alzheimer’s disease pathogenesis: Potential strategies to protect the LC against aging. Brain Research. 2019;1702:17–28. doi:10.1016/j.brainres.2017.12.027

7. Weinshenker D. Long Road to Ruin: Noradrenergic Dysfunction in Neurodegenerative Disease. Trends in Neurosciences. 2018;41(4):211–223. doi:10.1016/j.tins.2018.01.010

8. Mather M, Harley CW. The Locus Coeruleus: Essential for Maintaining Cognitive Function and the Aging Brain. Trends in Cognitive Sciences. 2016;20(3):214–226. doi:10.1016/j.tics.2016.01.001

9. Galgani A, Giorgi FS. Exploring the Role of Locus Coeruleus in Alzheimer’s Disease: a Comprehensive Update on MRI Studies and Implications. Curr Neurol Neurosci Rep. 2023;23(12):925–936. doi:10.1007/s11910-023-01324-9

10. Betts MJ, Cardenas-Blanco A, Kanowski M, Jessen F, Düzel E. In vivo MRI assessment of the human locus coeruleus along its rostrocaudal extent in young and older adults. NeuroImage. 2017;163:150–159. doi:10.1016/j.neuroimage.2017.09.042

11. Wearn A, Onuska KM, Shanks HRC, et al. Locus coeruleus degeneration is associated with cortical tau deposition and cognitive decline in older adults at familial risk of Alzheimer’s disease. Alzheimer’s & Dementia. 2026;22(4):e71427. doi:10.1002/alz.71427

12. Dahl MJ, Mather M, Werkle-Bergner M, et al. Locus coeruleus integrity is related to tau burden and memory loss in autosomal-dominant Alzheimer’s disease. Neurobiology of Aging. 2022;112:39–54. doi:10.1016/j.neurobiolaging.2021.11.006

13. Engels-Domínguez N, Riphagen JM, Van Egroo M, et al. Lower Locus Coeruleus Integrity Signals Elevated Entorhinal Tau and Clinical Progression in Asymptomatic Older Individuals. Annals of Neurology. 2024;96(4):650–661. doi:10.1002/ana.27022

14. Jacobs HIL, Becker JA, Kwong K, et al. Waning locus coeruleus integrity precedes cortical tau accrual in preclinical autosomal dominant Alzheimer’s disease. Alzheimer’s & Dementia. 2023;19(1):169–180. doi:10.1002/alz.12656

15. Bueichekú E, Diez I, Kim CM, et al. Spatiotemporal patterns of locus coeruleus integrity predict cortical tau and cognition. Nat Aging. 2024;4(5):625–637. doi:10.1038/s43587-024-00626-y

16. Sibahi A, Hosseini SA, Rahmouni N, et al. A longitudinal investigation of locus coeruleus norepinephrine system integrity and cognitive performance across the Alzheimer’s spectrum. J Cereb Blood Flow Metab. Published online June 5, 2026:271678X261460160. doi:10.1177/0271678X261460160

17. Jack CR, Andrews JS, Beach TG, et al. Revised criteria for diagnosis and staging of Alzheimer’s disease: Alzheimer’s Association Workgroup. Alzheimer’s & Dementia. 2024;20(8):5143–5169. doi:10.1002/alz.13859

18. Rabinovici GD, Knopman DS, Arbizu J, et al. Updated appropriate use criteria for amyloid and tau PET: A report from the Alzheimer’s Association and Society for Nuclear Medicine and Molecular Imaging Workgroup. Alzheimer’s & Dementia. 2025;21(1):e14338. doi:10.1002/alz.14338

19. Lesman-Segev OH, La Joie R, Iaccarino L, et al. Diagnostic Accuracy of Amyloid versus^18^ F-Fluorodeoxyglucose Positron Emission Tomography in AUTOPSY-CONFIRMED Dementia. Annals of Neurology. 2021;89(2):389–401. doi:10.1002/ana.25968

20. Montoliu-Gaya L, Salvadó G, Therriault J, et al. Plasma tau biomarkers for biological staging of Alzheimer’s disease. Nat Aging. 2025;5(11):2297–2308. doi:10.1038/s43587-025-00951-w

21. Wu H, Liu L, Zeng L. Advances in plasma biomarkers for the diagnosis of Alzheimer’s disease. Brain Research. 2026;1874:150138. doi:10.1016/j.brainres.2025.150138

22. Ashton NJ, Brum WS, Di Molfetta G, et al. Diagnostic Accuracy of a Plasma Phosphorylated Tau 217 Immunoassay for Alzheimer Disease Pathology. JAMA Neurol. 2024;81(3):255. doi:10.1001/jamaneurol.2023.5319

23. Aarsland D, Sunde AL, Tovar-Rios DA, et al. Prevalence of Alzheimer’s disease pathology in the community. Nature. 2026;650(8100):182–186. doi:10.1038/s41586-025-09841-y

24. Salvadó G, Janelidze S, Bali D, et al. Plasma Phosphorylated Tau 217 to Identify Preclinical Alzheimer Disease. JAMA Neurol. 2025;82(11):1122–1134. doi:10.1001/jamaneurol.2025.3217

25. Malek-Ahmadi M, Sharma S, Stipho F, et al. Plasma Phosphorylated Tau 217 and Amyloid Burden in Older Adults Without Cognitive Impairment: A Meta-Analysis. JAMA Neurol. 2026;83(1):13–19. doi:10.1001/jamaneurol.2025.4721

26. Janelidze S, Berron D, Smith R, et al. Associations of Plasma Phospho-Tau217 Levels With Tau Positron Emission Tomography in Early Alzheimer Disease. JAMA Neurol. 2021;78(2):1–8. doi:10.1001/jamaneurol.2020.4201

27. Mattsson-Carlgren N, Janelidze S, Bateman RJ, et al. Soluble P-tau217 reflects amyloid and tau pathology and mediates the association of amyloid with tau. EMBO Mol Med. 2021;13(6):EMMM202114022. doi:10.15252/emmm.202114022

28. Ferreira PCL, Bellaver B, Povala G, et al. Evaluating Plasma p-tau217 as an Endpoint for Alzheimer Disease Clinical Trials. Neurology. 2026;106(1):e214441. doi:10.1212/WNL.0000000000214441

29. Yang HS, Anzai JAU, Yau WYW, et al. Plasma phosphorylated tau 217 and longitudinal trajectories of Aβ, tau, and cognition in cognitively unimpaired older adults. Nat Commun. 2026;17(1):3188. doi:10.1038/s41467-026-71269-3

30. Wilson D, Khare M, Triana-Baltzer G, et al. Analytical and Clinical Validation of a High Accuracy Fully Automated Digital Immunoassay for Plasma Phospho-Tau 217 for Clinical Use in Detecting Amyloid Pathology. Neurology. Preprint posted online November 4, 2024. doi:10.1101/2024.10.31.24316186

31. Palmqvist S, Warmenhoven N, Anastasi F, et al. Plasma phospho-tau217 for Alzheimer’s disease diagnosis in primary and secondary care using a fully automated platform. Nat Med. 2025;31(6):2036–2043. doi:10.1038/s41591-025-03622-w

32. Galgani A, Mary A, Lombardo F, et al. Locus coeruleus integrity correlates with plasma soluble Axl levels in Alzheimer’s disease patients. Alzheimer’s & Dementia. 2025;21(7):e70434. doi:10.1002/alz.70434

33. David MCB, Kolanko MA, Parker TD, et al. Catecholaminergic nucleus integrity and Alzheimer’s pathology, symptoms, and progression. Alzheimer’s & Dementia. 2025;21(10):e70749. doi:10.1002/alz.70749

34. Van Egroo M, Riphagen JM, Ashton NJ, et al. Ultra-high field imaging, plasma markers and autopsy data uncover a specific rostral locus coeruleus vulnerability to hyperphosphorylated tau. Mol Psychiatry. 2023;28(6):2412–2422. doi:10.1038/s41380-023-02041-y

35. Sara SJ. The locus coeruleus and noradrenergic modulation of cognition. Nat Rev Neurosci. 2009;10(3):211–223. doi:10.1038/nrn2573

36. Dahl MJ, Bachman SL, Dutt S, et al. The integrity of dopaminergic and noradrenergic brain regions is associated with different aspects of late-life memory performance. Nat Aging. 2023;3(9):1128–1143. doi:10.1038/s43587-023-00469-z

37. Elman JA, Puckett OK, Beck A, et al. MRI-assessed locus coeruleus integrity is heritable and associated with multiple cognitive domains, mild cognitive impairment, and daytime dysfunction. Alzheimers Dement. 2021;17(6):1017–1025. doi:10.1002/alz.12261

38. Dahl MJ, Mather M, Düzel S, et al. Rostral locus coeruleus integrity is associated with better memory performance in older adults. Nat Hum Behav. 2019;3(11):1203–1214. doi:10.1038/s41562-019-0715-2

39. Pahl J, Prokopiou PC, Bueichekú E, et al. Locus coeruleus integrity and left frontoparietal connectivity provide resilience against attentional decline in preclinical alzheimer’s disease. Alzheimers Res Ther. 2024;16(1):119. doi:10.1186/s13195-024-01485-w

40. Lim YY, Kalinowski P, Pietrzak RH, et al. Association of β-Amyloid and Apolipoprotein E ε4 With Memory Decline in Preclinical Alzheimer Disease. JAMA Neurol. 2018;75(4):488–494. doi:10.1001/jamaneurol.2017.4325

41. Sperling RA, Aisen PS, Beckett LA, et al. Toward defining the preclinical stages of Alzheimer’s disease: Recommendations from the National Institute on Aging-Alzheimer’s Association workgroups on diagnostic guidelines for Alzheimer’s disease. Alzheimer’s & Dementia. 2011;7(3):280–292. doi:10.1016/j.jalz.2011.03.003

42. Dutt S, Bachman SL, Dahl MJ, et al. Locus coeruleus MRI contrast, cerebral perfusion, and plasma Alzheimer’s disease biomarkers in older adults. Neurobiol Aging. 2025;147:12–21. doi:10.1016/j.neurobiolaging.2024.11.008

43. Avants BB, Tustison NJ, Song G, Cook PA, Klein A, Gee JC. A reproducible evaluation of ANTs similarity metric performance in brain image registration. Neuroimage. 2011;54(3):2033–2044. doi:10.1016/j.neuroimage.2010.09.025

44. Liu KY, Marijatta F, Hämmerer D, Acosta-Cabronero J, Düzel E, Howard RJ. Magnetic resonance imaging of the human locus coeruleus: A systematic review. Neurosci Biobehav Rev. 2017;83:325–355. doi:10.1016/j.neubiorev.2017.10.023

45. Jak AJ, Bondi MW, Delano-Wood L, et al. Quantification of five neuropsychological approaches to defining mild cognitive impairment. Am J Geriatr Psychiatry. 2009;17(5):368–375. doi:10.1097/JGP.0b013e31819431d5

46. Kapoor A, Gaubert A, Marshall A, et al. Increased Levels of Circulating Angiogenic Cells and Signaling Proteins in Older Adults With Cerebral Small Vessel Disease. Front Aging Neurosci. 2021;13:711784. doi:10.3389/fnagi.2021.711784

47. Kapoor A, Dutt S, Engstrom AC, et al. Association of Medial Temporal Lobe Cerebrovascular Reactivity and Memory Function in Older Adults With and Without Cognitive Impairment. Neurology. 2025;104(1):e210210. doi:10.1212/WNL.0000000000210210

48. Aiken LS, West SG. Multiple Regression: Testing and Interpreting Interactions. Sage Publications, Inc; 1991:xi, 212.

49. Koops EA, Dutta J, Hanseeuw BJ, et al. Elevated locus coeruleus metabolism provides resilience against cognitive decline in preclinical Alzheimer’s disease. Alzheimer’s & Dementia. 2025;21(1):e14385. doi:10.1002/alz.14385

50. Wearn A, Onuska KM, Schmitz TW, Turner GR, Spreng RN. The neuromodulatory fragility hypothesis of Alzheimer’s disease pathogenesis. Alzheimers Dement. 2026;22(3):e71249. doi:10.1002/alz.71249

51. Milà-Alomà M, Ashton NJ, Shekari M, et al. Plasma p-tau231 and p-tau217 as state markers of amyloid-β pathology in preclinical Alzheimer’s disease. Nat Med. 2022;28(9):1797–1801. doi:10.1038/s41591-022-01925-w

52. Jacobs HIL. The monoaminergic systems as drivers of Alzheimer’s disease pathophysiology and symptomatology. Current Opinion in Neurology. 2026;39(2):192–199. doi:10.1097/WCO.0000000000001455

53. Jacobs HIL, Hämmerer D, Mather M, Poser BA, Rowe J. A call for a mechanistic-neutral nomenclature replacing “neuromelanin-sensitive” imaging. PsyArXiv. Preprint posted online September 9, 2026:94h6n_v1. Accessed October 3, 2026. https://osf.io/preprints/psyarxiv/94h6n_v1/

