## Supplemental Materials for "Locus coeruleus intensity as a marker of early-stage Alzheimer’s disease: associations with plasma pTau_217_ and memory"

**Supplemental Table S1** Results from logistic regression model examining the association between locus coeruleus intensity and plasma $pTau_{217}$ positivity

| | ***b*** | ***SE*** | **OR** | **95% CI** | ***P*** |
|---|---|---|---|---|---|
| Intercept | -8.69 | 2.05 | | | |
| LC intensity | 0.43 | 0.20 | 1.53 | 1.05, 2.31 | **.033** |
| Age | 0.10 | 0.03 | 1.11 | 1.05, 1.17 | < .001 |
| Sex | -0.33 | 0.41 | 0.72 | 0.31, 1.58 | .419 |
| *APOE* ε4 carrier status | 1.21 | 0.40 | 3.35 | 1.55, 7.52 | .003 |

*Note.* Unstandardized estimates, standard errors, and 95% CIs are presented. *APOE* ε4 carrier status was dummy-coded such that 0 = noncarrier and 1 = carrier and sex was dummy-coded such that 0 = male and 1 = female. LC intensity was standardized (*z*-scored) and reverse-coded such that a one-unit increase represents a 1-*SD* decrease in LC intensity; therefore, the odds ratio represents the change in odds of plasma $pTau_{217}$ positivity associated with a 1-*SD* decrease in LC intensity. Plasma $pTau_{217}$ positivity was defined as concentrations > 0.42 pg/mL.

Abbreviations: *APOE* ε4, apolipoprotein E ε4; *b*, unstandardized regression coefficient; CI, confidence interval; LC, locus coeruleus; OR, odds ratio; $pTau_{217}$, phosphorylated tau 217; *SD*, standard deviation; *SE*, standard error.

**Supplemental Table S2** Results from linear regression model examining the association between locus coeruleus intensity and plasma $pTau_{217}$ concentration, adjusted for cognitive status

| | ***b*** | ***SE*** | **95% CI** | ***P*** |
|---|---|---|---|---|
| Intercept | -0.43 | 0.17 | | |
| LC intensity | -1.01 | 0.46 | -1.91, -0.10 | **.029** |
| Age | 0.01 | < 0.01 | 0.01, 0.02 | < .001 |
| Sex | 0.01 | 0.04 | -0.06, 0.07 | .887 |
| *APOE* ε4 carrier status | 0.14 | 0.03 | 0.07, 0.21 | < .001 |
| Cognitive status | 0.03 | 0.04 | -0.06, 0.11 | .533 |

*Note.* Unstandardized estimates, standard errors, and 95% CIs are presented. *APOE* ε4 carrier status was dummy-coded such that 0 = noncarrier and 1 = carrier and sex was dummy-coded such that 0 = male and 1 = female. Cognitive status was dummy coded such that 0 = cognitively unimpaired and 1 = probable mild cognitive impairment.

Abbreviations: *APOE* ε4, apolipoprotein E ε4; *b*, unstandardized regression coefficient; CI, confidence interval; LC, locus coeruleus; $pTau_{217}$, phosphorylated tau 217; *SE*, standard error.

**Supplemental Table S3** Results from logistic regression model examining the association between locus coeruleus intensity and plasma $pTau_{217}$ positivity, adjusted for cognitive status

| | ***b*** | ***SE*** | **OR** | **95% CI** | ***P*** |
|---|---|---|---|---|---|
| Intercept | -8.72 | 2.08 | | | |
| LC intensity | 0.41 | 0.20 | 1.50 | 1.02, 2.27 | **.044** |
| Age | 0.10 | 0.03 | 1.11 | 1.05, 1.17 | < .001 |
| Sex | -0.33 | 0.42 | 0.72 | 0.31, 1.61 | .434 |
| *APOE* ε4 carrier status | 1.27 | 0.41 | 3.54 | 1.63, 8.05 | .002 |
| Cognitive status | 0.42 | 0.48 | 1.53 | 0.59, 3.89 | .376 |

*Note.* Unstandardized estimates, standard errors, and 95% CIs are presented. *APOE* ε4 carrier status was dummy-coded such that 0 = noncarrier and 1 = carrier and sex was dummy-coded such that 0 = male and 1 = female. LC intensity was standardized (*z*-scored) and reverse-coded such that a one-unit increase represents a 1-*SD* decrease in LC intensity; therefore, the odds ratio represents the change in odds of plasma $pTau_{217}$ positivity associated with a 1-*SD* decrease in LC intensity. Cognitive status was dummy coded such that 0 = cognitively unimpaired and 1 = probable mild cognitive impairment. Plasma $pTau_{217}$ positivity was defined as concentrations > 0.42 pg/mL.

Abbreviations: *APOE* ε4, apolipoprotein E ε4; *b*, unstandardized regression coefficient; CI, confidence interval; LC, locus coeruleus; OR, odds ratio; $pTau_{217}$, phosphorylated tau 217; *SD*, standard deviation; *SE*, standard error.

**Supplemental Table S4** Results from multiple linear regression model examining the interaction between locus coeruleus intensity and plasma $pTau_{217}$ positivity in relation to memory scores

| | ***b*** | ***SE*** | **95% CI** | ***P*** |
|---|---|---|---|---|
| Intercept | -0.27 | 0.85 | | |
| LC intensity | 5.75 | 2.41 | 0.98, 10.52 | .019 |
| Plasma $pTau_{217}$ positivity | 0.37 | 0.25 | -0.14, 0.87 | .151 |
| Age | -0.03 | 0.01 | -0.05, -0.01 | .011 |
| Sex | -0.39 | 0.15 | -0.69, -0.09 | .012 |
| Education | 0.12 | 0.03 | 0.05, 0.18 | < .001 |
| *APOE* ε4 carrier status | 0.21 | 0.15 | -0.09, 0.50 | .171 |
| LC intensity × plasma $pTau_{217}$ positivity | -10.21 | 4.11 | -18.34, -2.07 | **.014** |

*Note.* Unstandardized estimates, standard errors, and 95% CIs are presented. LC intensity was mean-centered prior to analysis. *APOE* ε4 carrier status was dummy-coded such that 0 = noncarrier and 1 = carrier and sex was dummy-coded such that 0 = male and 1 = female. Plasma $pTau_{217}$ positivity was defined as concentrations > 0.42 pg/mL and pTau217 status was coded such that 0 = negative and 1 = positive.

Abbreviations: *APOE* ε4, apolipoprotein E ε4; *b*, unstandardized regression coefficient; CI, confidence interval; LC, locus coeruleus; $pTau_{217}$, phosphorylated tau 217; *SE*, standard error.

**Supplemental Table S5** Results from multiple linear regression model examining the interaction between locus coeruleus intensity and plasma $pTau_{217}$ in relation to memory scores, adjusted for cognitive status

| | ***b*** | ***SE*** | **95% CI** | ***P*** |
|---|---|---|---|---|
| Intercept | 0.32 | 0.72 | | |
| LC intensity | 2.06 | 1.66 | -1.23, 5.35 | .218 |
| Plasma $pTau_{217}$ | -0.43 | 0.31 | -1.04, 0.18 | .166 |
| Age | -0.02 | 0.01 | -0.04, -0.01 | .006 |
| Sex | -0.24 | 0.13 | -0.49, 0.01 | .064 |
| Education | 0.10 | 0.03 | 0.05, 0.15 | < .001 |
| *APOE* ε4 carrier status | 0.12 | 0.13 | -0.13, 0.36 | .360 |
| Cognitive status | -1.09 | 0.14 | -1.37, -0.81 | < .001 |
| LC intensity × plasma $pTau_{217}$ | -18.73 | 6.00 | -30.63, -6.84 | **.002** |

*Note.* Unstandardized estimates, standard errors, and 95% CIs are presented. LC intensity and plasma $pTau_{217}$ were mean-centered prior to analysis. *APOE* ε4 carrier status was dummy-coded such that 0 = noncarrier and 1 = carrier and sex was dummy-coded such that 0 = male and 1 = female. Cognitive status was dummy-coded such that 0 = cognitively unimpaired and 1 = probable mild cognitive impairment.

Abbreviations: *APOE* ε4, apolipoprotein E ε4; *b*, unstandardized regression coefficient; CI, confidence interval; LC, locus coeruleus; $pTau_{217}$, phosphorylated tau 217; *SE*, standard error.

**Supplemental Table S6** Results from multiple linear regression model examining the interaction between locus coeruleus intensity and plasma $pTau_{217}$ positivity in relation to memory scores, adjusted for cognitive status

| | ***b*** | ***SE*** | **95% CI** | ***P*** |
|---|---|---|---|---|
| Intercept | 0.70 | 0.69 | | |
| LC intensity | 4.24 | 2.01 | 0.26, 8.21 | .037 |
| Plasma $pTau_{217}$ positivity | -0.11 | 0.15 | -0.40, 0.19 | .465 |
| Age | -0.03 | 0.01 | -0.04, -0.01 | .002 |
| Sex | -0.26 | 0.13 | -0.52, -0.01 | .042 |
| Education | 0.09 | 0.03 | 0.04, 0.15 | < .001 |
| *APOE* ε4 carrier status | 0.08 | 0.13 | -0.17, 0.33 | .509 |
| Cognitive status | -1.08 | 0.14 | -1.36, -0.79 | < .001 |
| LC intensity × plasma $pTau_{217}$ positivity | -8.81 | 3.41 | -15.56, -2.06 | **.011** |

*Note.* Unstandardized estimates, standard errors, and 95% CIs are presented. LC intensity was mean-centered prior to analysis. *APOE* ε4 carrier status was dummy-coded such that 0 = noncarrier and 1 = carrier and sex was dummy-coded such that 0 = male and 1 = female. Cognitive status was dummy-coded such that 0 = cognitively unimpaired and 1 = probable mild cognitive impairment. Plasma $pTau_{217}$ positivity was defined as concentrations > 0.42 pg/mL and $pTau_{217}$ status was coded 0 = negative and 1 = positive.

Abbreviations: *APOE* ε4, apolipoprotein E ε4; *b*, unstandardized regression coefficient; CI, confidence interval; LC, locus coeruleus; $pTau_{217}$, phosphorylated tau 217; *SE*, standard error.
